# Vaccine Breakthrough Infection with the SARS-CoV-2 Delta or Omicron (BA.1) Variant Leads to Distinct Profiles of Neutralizing Antibody Responses

**DOI:** 10.1101/2022.03.02.22271731

**Authors:** Michael S. Seaman, Mark J. Siedner, Julie Boucau, Christy L. Lavine, Fadi Ghantous, May Y. Liew, Josh Mathews, Arshdeep Singh, Caitlin Marino, James Regan, Rockib Uddin, Manish C. Choudhary, James P. Flynn, Geoffrey Chen, Ashley M. Stuckwisch, Taryn Lipiner, Autumn Kittilson, Meghan Melberg, Rebecca F. Gilbert, Zahra Reynolds, Surabhi L. Iyer, Grace C. Chamberlin, Tammy D. Vyas, Jatin M. Vyas, Marcia B. Goldberg, Jeremy Luban, Jonathan Z. Li, Amy K. Barczak, Jacob E. Lemieux

## Abstract

There is increasing evidence that the risk of SARS-CoV-2 infection among vaccinated individuals is variant-specific, suggesting that protective immunity against SARS-CoV-2 may differ by variant. We enrolled vaccinated (n = 39) and unvaccinated (n = 11) individuals with acute, symptomatic SARS-CoV-2 Delta or Omicron infection and performed SARS-CoV-2 viral load quantification, whole-genome sequencing, and variant-specific antibody characterization at the time of acute illness and convalescence. Viral load at the time of infection was inversely correlated with antibody binding and neutralizing antibody responses. Increases in antibody titers and neutralizing activity occurred at convalescence in a variant-specific manner. Across all variants tested, convalescent neutralization titers in unvaccinated individuals were markedly lower than in vaccinated individuals. For individuals infected with the Delta variant, neutralizing antibody responses were weakest against BA.2, whereas infection with Omicron BA.1 variant generated a broader response against all tested variants, including BA.2.

## Introduction

Breakthrough infection following SARS-CoV-2 vaccination has been observed since the rollout of vaccines^1,2^ and has been associated with specific variants including Beta^3^, Delta^4^, and Omicron^5^. The predisposing factors to breakthrough infection and the consequences of breakthrough infection for SARS-CoV-2 immunity are poorly understood. Both host- and viral-factors have been implicated. Immunocompromised patients mount poor immune responses after vaccination^6^ and are at high risk of infection despite vaccination^7^. Further, early studies demonstrated breakthrough infections despite neutralizing activity of patient serum at the time of infection^2^. The association with specific SARS-CoV-2 variants suggests that viral factors also play a role. Because T-cell reactivity to SARS-CoV-2 peptides appears largely preserved across variants^8,9^, but neutralizing activity of antibodies is markedly decreased to several variants, especially Omicron^10–12^, we hypothesized that antibody-specific responses differ at the time of vaccine breakthrough infection in a variant specific manner. We enrolled a cohort of ambulatory individuals with symptomatic breakthrough infection and characterized the viral genotype, viral load, and host antibody response at the time of breakthrough infection and after recovery.

## Methods

### Study recruitment

This study was approved by the Mass General Brigham institutional review board under protocol 2021P000812. Symptomatic adult outpatients diagnosed with COVID-19 were recruited based on a positive SARS-CoV-2 test. Informed consent was obtained from all participants prior to initiating study procedures. We conducted home visits. Phlebotomy was performed at the initial visit and at subsequent visit approximately 14 days later. Anterior nasal swabs were collected three times weekly at home visits until negative PCR testing and stored in viral transport medium. Specimens were transported to the laboratory within four hours of collection. Viral transport medium containing anterior nasal swabs was aliquoted, and stored at −80 C until testing. To isolate serum, blood was collected into BD red-top (anticoagulant-free) vacutainer tubes and processed according to manufacturer instructions. Serum was stored in aliquots at −80 C until processing.

### Laboratory methods

Viral genotype was determined using Spike gene sequencing and whole-genome sequencing as previously described ^13^. Viral load quantification and viral culture were also performed as previously described ^13^. Sequence data were submitted to Genbank under accession number PRJNA759255. ELISA assays were performed using the Elecsys Anti-SARS-CoV-2 ELISA assays for Spike and Nucleocapsid antibodies as per manufacturer instructions (presumed wild type). Concentrations exceeding 25,000 U/mL were set to the assay maximum of 25,000 and non-parametric tests in paired comparisons were used to account for this. Pseudovirus neutralization assays were performed as previously described ^14^. Serum samples were tested in duplicate using a primary 1:20 dilution with 3-fold or 5-fold titration series. Patients were enrolled in the study as soon as possible after diagnosis of COVID-19. We defined vaccinated patients as having completed a full primary immunization series, either two doses of mrna-1273 or bnt162b2 or a single dose of Ad26.COV.S at least 14 days prior to enrollment. We defined boosted patients as having completed three doses of either mrna-1273 or bnt162b2 at least 14 days prior to enrollment.

### Statistical analysis

Statistical analysis was performed using R v4.0.1^15^ and visualized using ggplot2^16^ and the ggpubr package. Wilcoxon rank-sum tests to compare the medians between two groups. Kruskal-Wallis tests were used to compare the medians of multiple groups. For survival analysis, we used the Kaplan-Meyer method to estimate the survivor function for time to conversion of negative PCR.

## Results

We enrolled 50 ambulatory individuals with symptomatic SARS-CoV-2 infection. The clinical characteristics of this cohort are shown in Table 1. The cohort included patients infected with Delta (n = 19) and Omicron (n = 31) variants, and individuals who were unvaccinated (n=11), vaccinated (n=24), and boosted (n=15). All patients were outpatients. We collected a nasal swab and drew blood at the time of acute infection (median 4 days after the onset of symptoms or positive PCR test, range 2-10 days) and again at convalescence (median 17 days after the onset of symptoms or positive PCR, range 14 - 24 days). We measured SARS-CoV-2 viral load, performed whole-genome and S gene sequencing, quantified total Spike- and Nucleocapsid-antibodies using ELISA, and performed pseudovirus neutralization assays.

**Table 1:** Cohort characteristics Summary of demographic and clinical characteristics of study participants. P-values represent comparison of characteristics between participants with Delta versus Omicron variant infections from chi-squared exact tests for categorical variables and rank-sum non-parametric testing for continuous variables.

|  | **Delta variant**<br>**infection (n=19)** | **Omicron variant**<br>**infection (n=31)** | **P-value** |
| --- | --- | --- | --- |
| Female, n (%) | 14 (75%) | 21 (67%) | 0.75 |
| Age, median (IQR) | 41 (35-55) | 44 (30-57) | 0.75 |
| Vaccination status, n (%) |  |  | 0.19 |
| Unvaccinated | 4 (21%) | 7 (24%) |  |
| Vaccinated | 13 (68%) | 14 (45%) |  |
| Boosted | 2 (11%) | 10 (32%) |  |
| Days since last vaccination,<br>median (IQR) | 234 (65-255) | 130 (50-251) | 0.43 |
| Symptomatic infection, n (%) | 18 (95%) | 31 (100%) | >0.99 |

IQR: Interquartile range

We first compared spike-specific antibody levels by vaccination status. All vaccinated patients had high levels of Spike-specific antibodies at the time of breakthrough infection (Figure S1), with a significant increase in antibody binding titers resulting from breakthrough infection for vaccinated individuals (Figure S1; p < 0.001, paired Wilcoxon rank-sum test). Nasal viral load at the time of infection correlated inversely with anti-Spike antibody levels at the time of acute infection (Figure 1A; r = −0.49, p = 0.001). Assessing baseline neutralization activity against individual variants, we found that neutralizing antibody titers against variants also correlated inversely with viral load at the time of breakthrough. This relationship was more variable than for binding antibody titers and was not significant (Figure 1B; r = −0.18, p = 0.34 for BA.1; r = −0.048, p = 0.86 for Delta). In addition, the nasal swabs from breakthrough infections with lower anti-Spike antibody titers were more likely to grow in culture (Figure 1C, p< 0.05 Wilcoxon rank-sum test), and individuals with higher acute anti-Spike antibody titers (by tertile) converted faster to negative PCR (Figure 1D, p < 0.05, log rank test).

**Figure 1:**
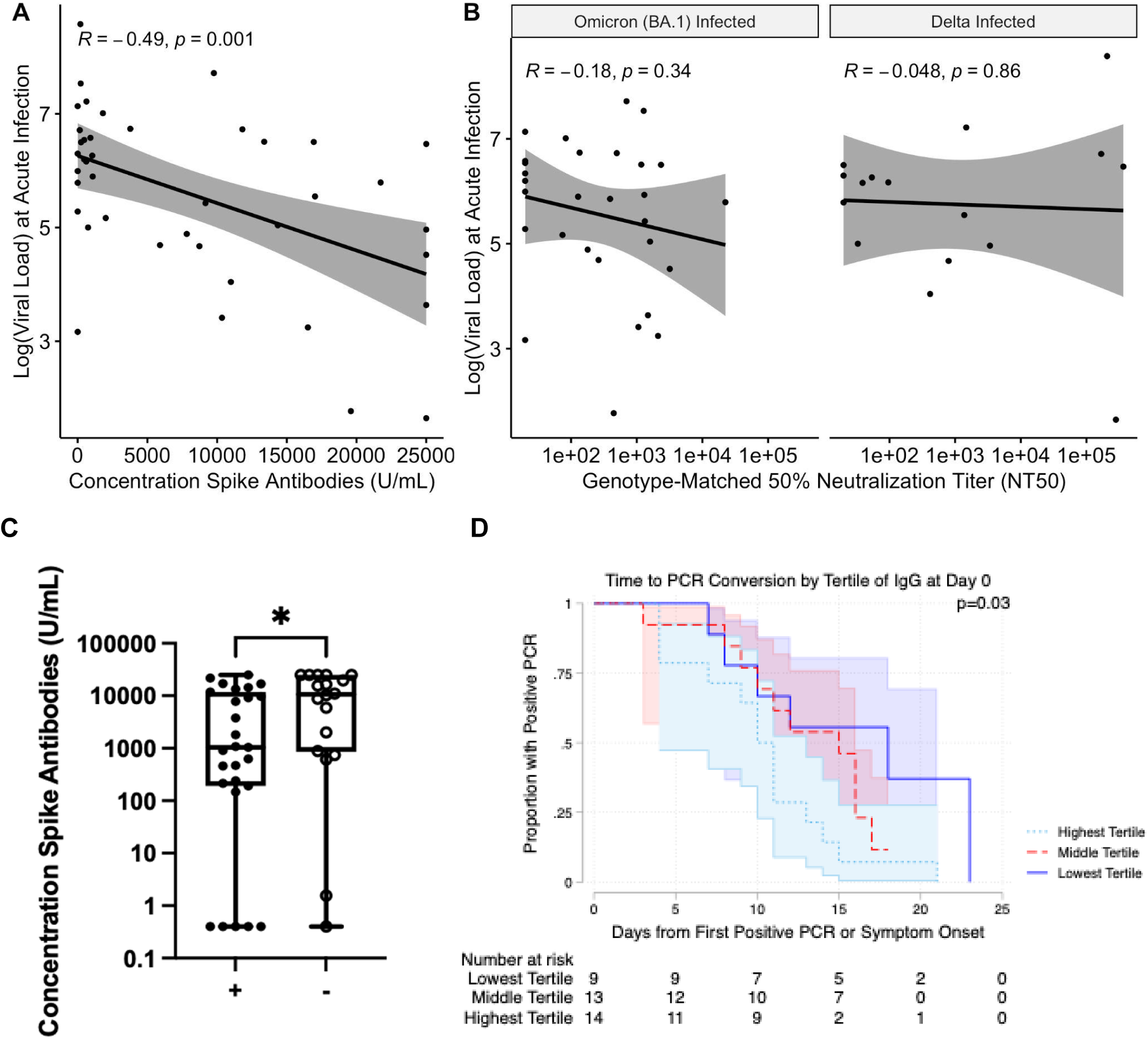
**A**. The logarithm of viral load at the time of infection vs the concentration of anti-Spike antibodies at the time of breakthrough. A regression line with standard error is shown, with the Pearson correlation coefficient and corresponding p value. **B**. Genotype-matched neutralizing antibody titers. A regression line with standard error is shown, with the Pearson correlation coefficient and corresponding p value. **C**. Anti-Spike antibody concentration at the initial study visit for culture-positive cases (+) and culture negative cases (−). Significance according to an unpaired Wilcoxon rank-sum test is shown. * - p < 0.05. **D**. Kaplan-Meier curves for time to PCR conversion by tertile of anti-Spike responses. The P-value represents log-rank testing comparing the subgroups.

We next examined differences in neutralization activity across timepoints and variants. As expected, neutralizing antibody responses against all the infecting variants increased significantly between acute infection and convalescence (Figure S2A and S3; Delta-infected against Delta pseudovirus, p < 0.001; BA.1-infected against BA.1 pseudovirus, p < 0.0001, paired Wilcoxon rank-sum test). Notably, infection also produced substantial increases in cross-neutralization (Figure S2A and S3; Delta-infected against BA.1 pseudovirus, p < 0.01, BA.1-infected against Delta pseudovirus, p < 0.001, paired Wilcoxon rank-sum test). Yet this effect was inconsistent: several participants, including 1 vaccinated individual infected with Delta, had no detectable neutralizing activity against BA.1 or BA.2 at convalescence. Overall, BA.2 pseudovirus was the most resistant to neutralization by convalescent serum: Among Delta-infected patients at convalescence, there was a 11.2-fold reduction in geometric mean titer against BA.2 as compared to Delta. Among BA.1-infected patients at convalescence, there was a 1.8-fold reduction in BA.2 titers as compared to BA.1. Convalescent titers and neutralizing activity were markedly lower for individuals who were unvaccinated at the time of infection, compared to those who had been previously vaccinated or boosted (Figure S4). Compared to boosted or vaccinated patients, unvaccinated patients had 38-fold lower neutralization titers against BA.2 (p < 0.0001, Wilcoxon rank-sum test), 31-fold lower against BA.1 (p < 0.001, Wilcoxon rank-sum test), 33-fold lower against D614G (p < 0.01, Wilcoxon rank-sum test), and 25-fold lower (p < 0.05, Wilcoxon rank-sum test) against Delta.

Variant-specific neutralizing activity differed by infecting variant, with Delta-infected patients developing a response that was more specific to WT and Delta variants (Figure 2). Among Delta-infected patients, neutralizing antibody titers differed significantly among variants at both acute (p = < 0.01, Kruskal-Wallis test) and convalescent timepoints (p < 0.01; Kruskal-Wallis test). In contrast, Omicron-infected patients had neutralizing activity that did not differ significantly among groups (p > 0.05 for acute and convalescent timepoints, Kruskal-Wallis test). Despite a response in Delta-infected individuals that was more specific toward WT and Delta variants, titers in that group still increased against BA.1 and BA.2, indicating an expansion of the antibody repertoire against variants not yet encountered. Convalescent serum from BA.1-infected participants demonstrated significantly higher neutralization titers against BA.1 and BA.2 compared to convalescent serum from Delta-infected participants (Figure S2B; p < 0.05, p < 0.05, Wilcoxon rank-sum test), while maintaining comparable neutralization titers against Delta pseudoviruses (Figure S2B).

**Figure 2:**
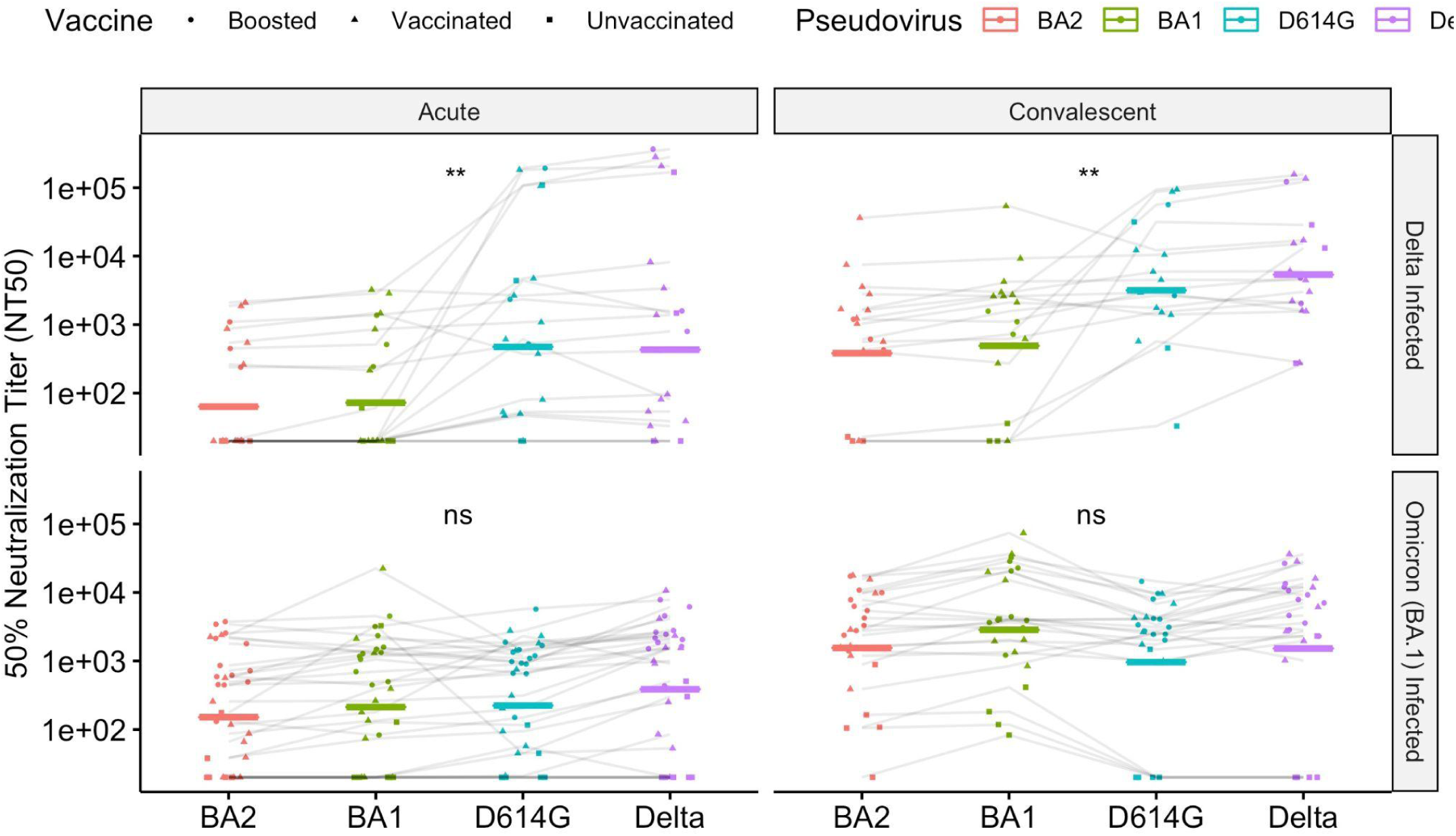
Neutralizing antibody responses, as measured by NT50, against a panel of pseudoviruses. The geometric mean is shown with a bar. Patients infected with Delta variants are shown in the top panel. Patients infected with the Omicron (BA.1) variant are shown in the bottom panel. Significance test of medians (Kruskal-Wallis) is shown. ns: p > 0.05; ** - p < 0.01.

We applied principal component analysis to characterize further the space of neutralizing antibody responses across variants (Figure 3). The first principal component corresponded roughly to strength of immunity, with positive values of the first principal component (PC1, variance explained 67%) corresponding to stronger neutralization responses for all variants tested (Figure 3, variant-specific loadings are plotted as arrows). As a result, unvaccinated cases separated from boosted cases by PC1 alone (Figure 3A). The second principal component (PC2, variance explained 19%) correlated with the infecting variant, with BA.1-infected cases clustering together. There was substantial overlap in the responses of Delta and Omicron-infected patients, particularly among the vaccinated and/or boosted, consistent with the broad-neutralizing responses observed for most patients. Taken together, these results suggest that vaccination status is the primary driver of differences in antibody responses across infected individuals, and that the infecting variant makes a modest additional contribution.

**Figure 3:**
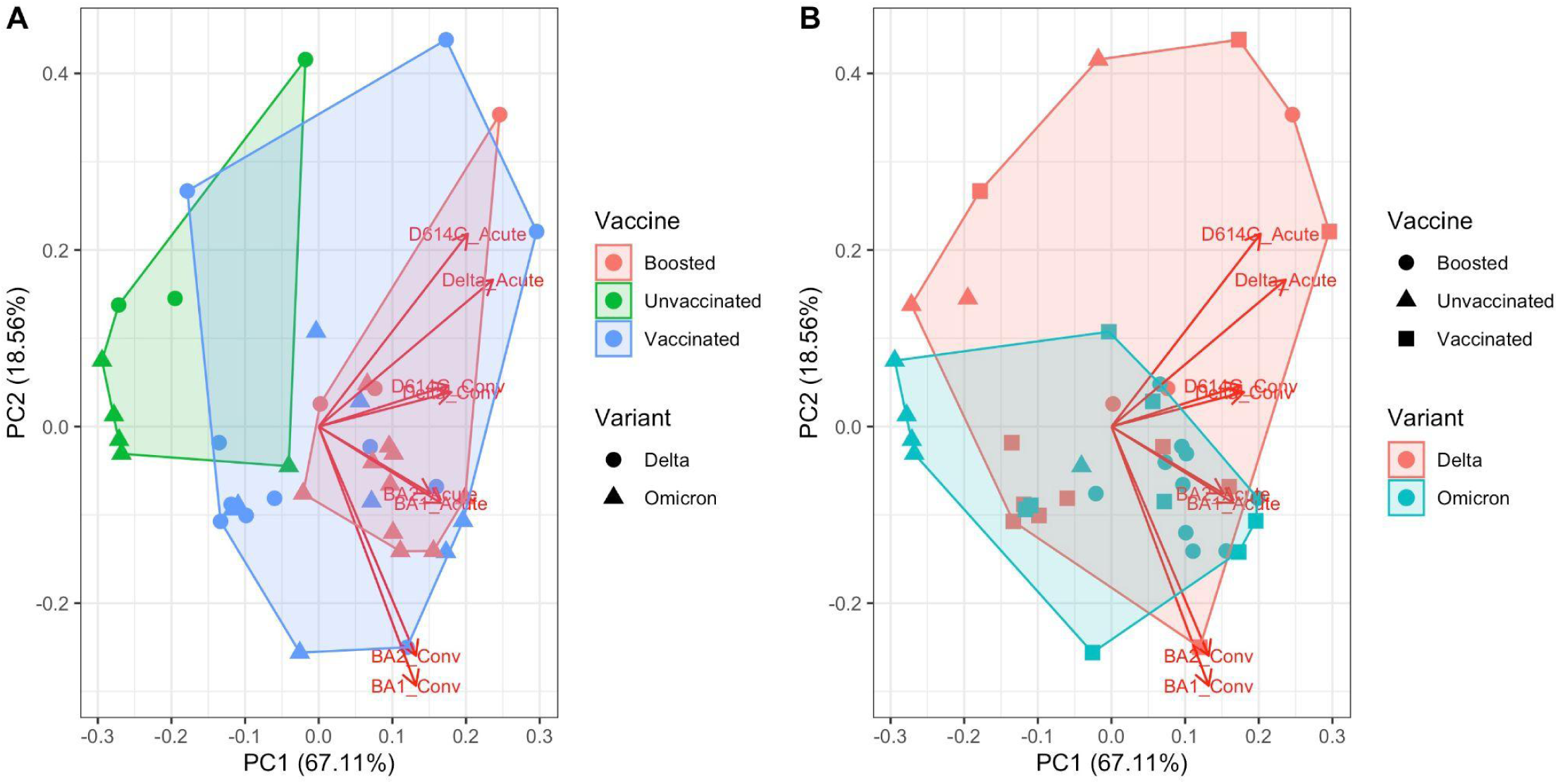
Principal component analysis of NT50 titers. The loadings are plotted as arrows. **A**. The convex hull of the clusters according to vaccination status is shaded. Point shape denotes the infecting variant. **B**. The convex hull of the clusters according to the infecting variant is shaded. Point shape denotes vaccination status at the time of infection.

## Conclusions

Breakthrough infection among vaccinated persons is an increasingly common presentation of SARS-CoV-2 infection with novel variants. Previous studies have linked both host and viral factors to the risk of reinfection, but much remains to be learned about the determinants of protection from SARS-CoV-2 infection. Understanding the breadth and potency of immunity against SARS-CoV-2 after vaccination in a variant-specific manner is critical for defining the limitations of the current generation of SARS-CoV-2 vaccines and developing broadly-neutralizing vaccine approaches in the future. We show here that antibody binding and neutralizing antibody titers correlate inversely with viral load at the time of breakthrough infection. Compared to infection with the Delta variant, we found that BA.1 infection induced neutralizing antibodies with greater breadth, including against Delta, BA.1 and BA.2 pseudoviruses.

Neutralization titers have previously been inversely linked to viral load in COVID-19 among unvaccinated patients ^17^. It is notable that this relationship also holds true in a vaccine-breakthrough cohort. We also show that breakthrough infections among vaccinated patients infected with Delta and Omicron variants induce high-titer neutralizing antibody responses, with the broadest responses being observed after Omicron infection. A subset of Delta-infected individuals showed no neutralizing antibody activity against Omicron variants.

Our results are directly relevant to future SARS-CoV-2 vaccine development. The increased breadth of neutralizing antibody repertoire seen in BA.1-infected, vaccinated individuals compared to Delta-infected, vaccinated individuals suggests that booster regimens that span the antigenic landscape of circulating SARS-CoV-2 genetic variation may have potential as one component of a future vaccination strategy to elicit broadly neutralizing antibody responses. Observations of responses in humans naturally-infected with different variants complement controlled studies in animal models. Although infection is not directly comparable to vaccination, our results are consistent with the observed enhancement of neutralization activity after boosting an mRNA-1273 primary series with mRNA-1273-Omicron^18^. Non-human primate studies of the mRNA-1273 booster, however, show no differences in neutralization response^19^. Defining the strategies that elicit durable immunity against current and future variants is an important area of SARS-CoV-2 research.

Our findings are also potentially relevant to SARS-CoV-2 epidemiology. The Omicron variant has overtaken the globe faster than any previous SARS-CoV-2 variant ^20^. Its increased fitness appears linked, at least in part, to its extensive antibody evasion properties and ability to cause vaccine breakthrough infections^21^ and reinfections^5^. Initially, most Omicron cases were caused by BA.1, but the BA.2 variant appears to have a higher growth rate in most populations and is replacing BA.1 as the dominant variant in many locations ^22,23^. Among the variants tested here, neutralization titers were lowest against BA.2. After BA.1 infection, we found a modest 1.8-fold decrease in geometric mean in BA.2 neutralization compared to BA.1 neutralization, roughly concordant with other recently reported data^24,25^. Nevertheless, because estimates are consistent in showing a 30-40% growth advantage per viral generation of BA.2 over BA.1 ^22,23^, and BA.1 appears to derive much of its fitness advantage from antibody escape^22^, these modest differences in neutralization titers may be epidemiologically significant.

The weak neutralizing antibody responses following infection in unvaccinated persons suggests that convalescent immunity alone, in the absence of vaccination, is unlikely to be sufficient to afford protection against future variants. In summary, our results highlight the increase in neutralizing activity following variant-specific breakthrough infection and emphasize its breadth across variants, particularly among vaccinated persons infected with Omicron BA.1. However, our findings also suggest that broad swaths of the population remain susceptible to circulating variants and underscore the importance of vaccination efforts.

## Data Availability

Sequence data were submitted to Genbank under accession number PRJNA759255

## Acknowledgements

The authors gratefully acknowledge Nicole-Doria Rose (NIAID) for sharing BA.1 and BA.2 plasmids. The authors would like to thank the MassCPR Executive Committee and administrative team, the MGH Clinical Microbiology lab, MGH Clinical Core lab, and the patients who participated in the study.

## Competing interests

none

**Figure S1:**
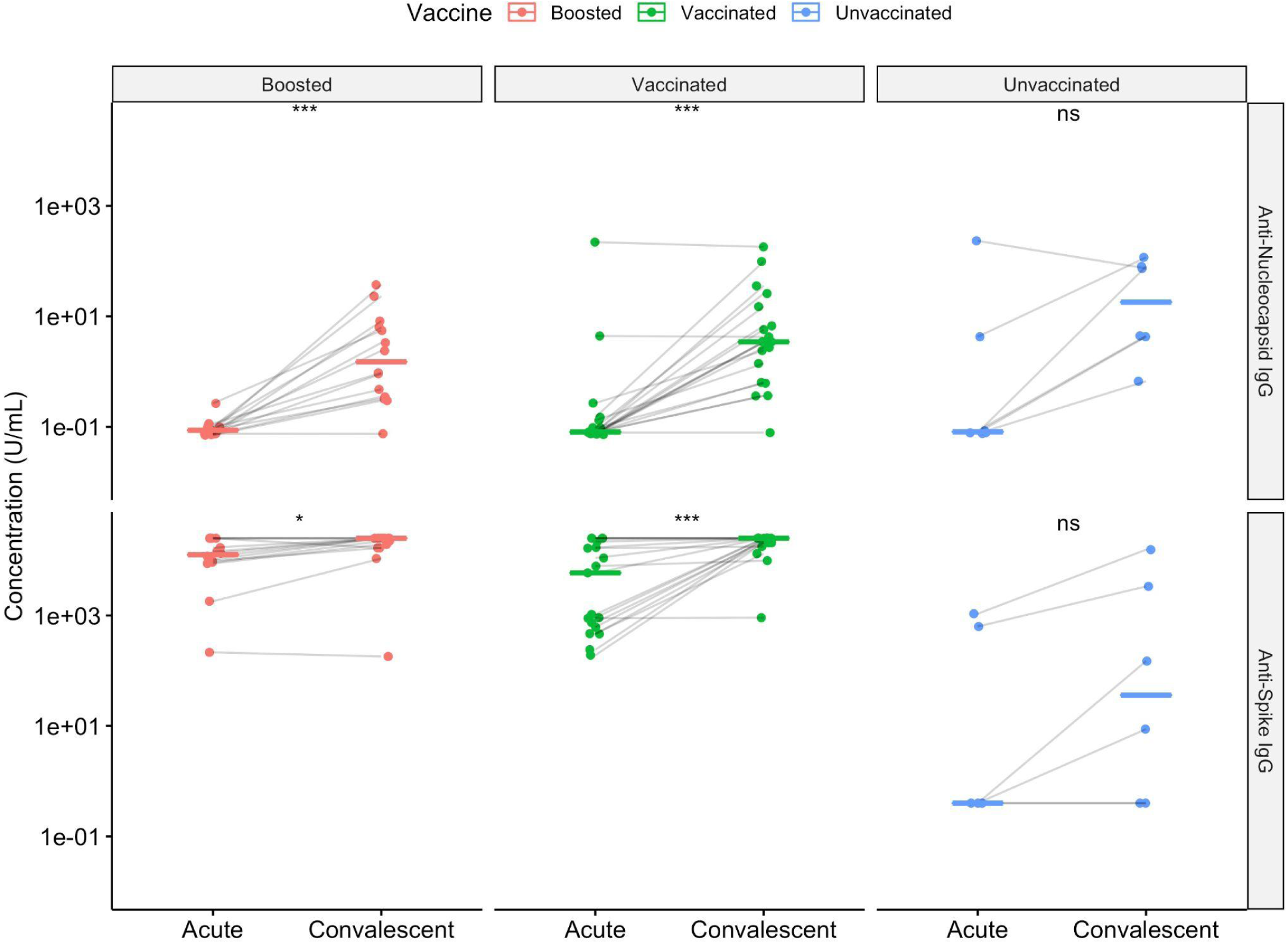
**A**. Antibody titers as measured by ELISA against Spike and Nucleocapsid protein at the time of acute infection and convalescence. Significance from a paired Wilcoxon test is shown. **B**. Antibody titers against Spike and Nucleocapsid protein by vaccination status. Significance from an unpaired Wilcoxon test is shown. ns: p > 0.05; * - p < 0.05; *** - p < 0.001; **** - p < 0.0001.

**Figure S2:**
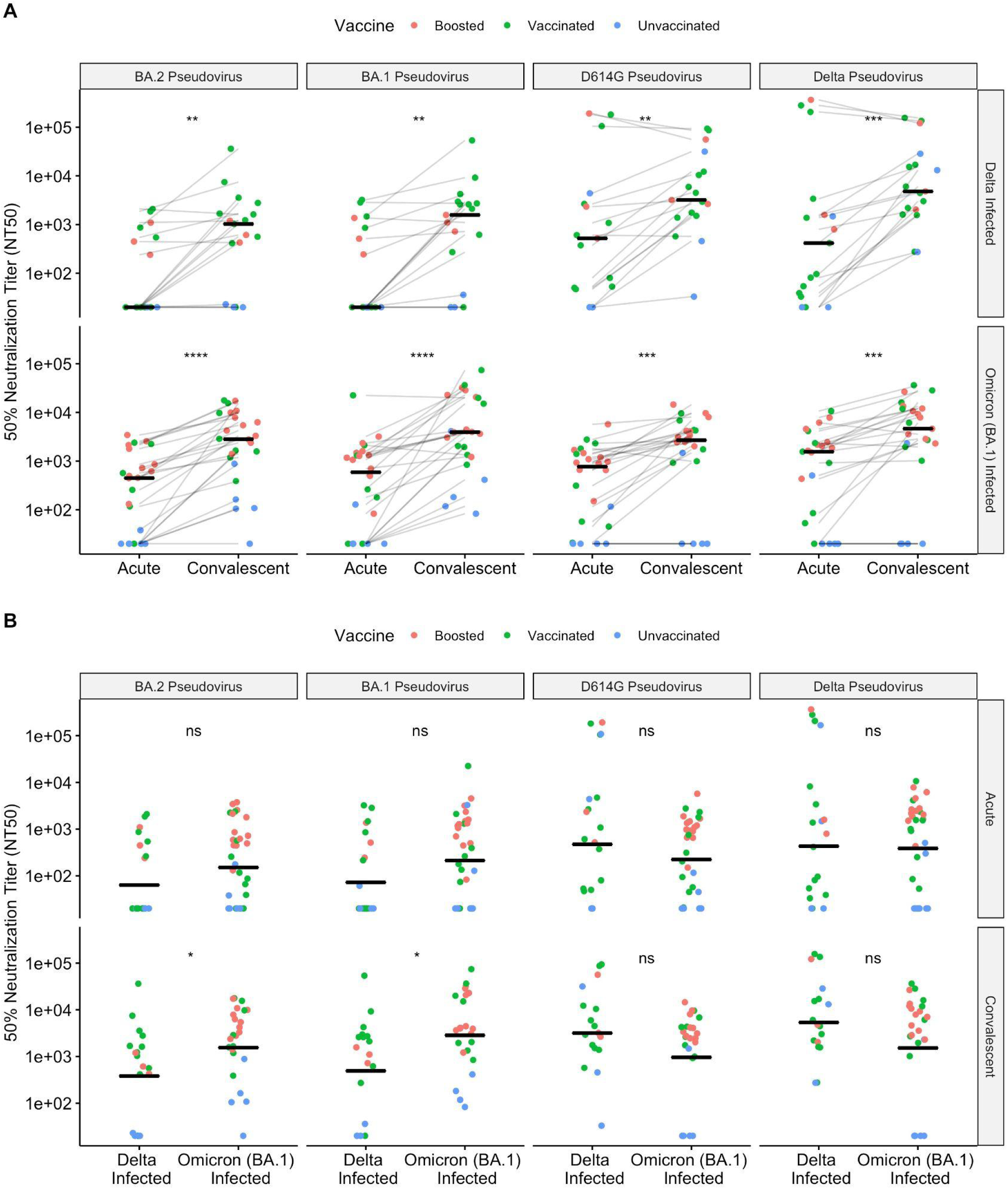
**A**. Neutralizing antibody responses against a panel of pseudoviruses, as measured by the 50% neutralizing antibody titer (NT50), at the time of acute infection and convalescence against a panel of pseudoviruses. Significance from a paired Wilcoxon test is shown. **B**. Neutralizing antibody responses by the infecting variant. Significance from an unpaired Wilcoxon test is shown. ns - p > 0.05; * - p < 0.05; ** - p < 0.01; *** - p < 0.001; **** - p < 0.0001.

**Figure S3:**
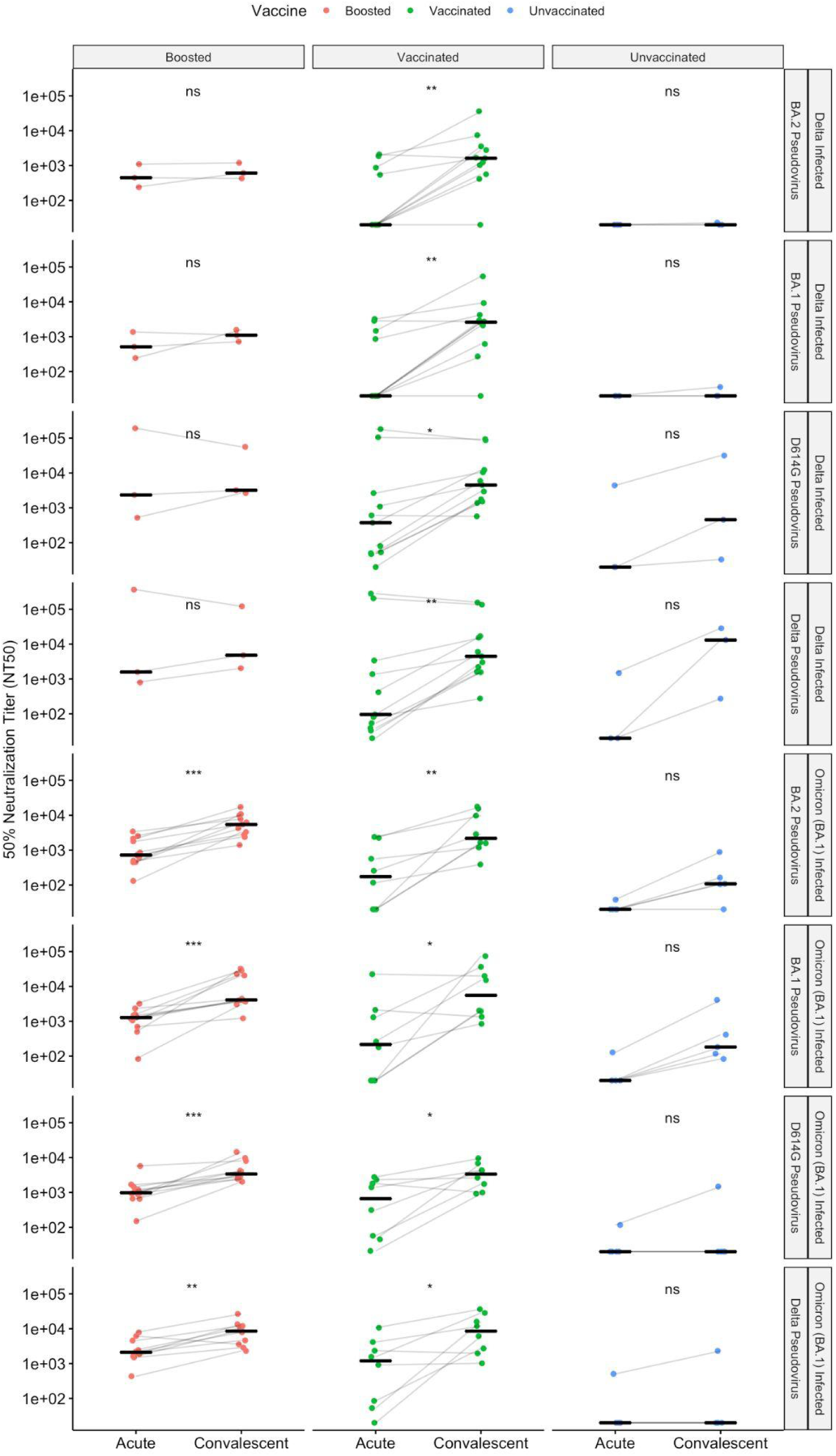
Neutralizing antibody responses, as measured by the 50% neutralizing antibody titer (NT50), at the time of acute infection and convalescence against a panel of pseudoviruses. Responses are stratified by the genotype of the infecting variant, the genotype of the pseudovirus, and the vaccination status of the individual. Significance from a paired Wilcoxon test is shown. ns: p > 0.05; * - p < 0.05; ** - p < 0.01; *** - p < 0.001.

**Figure S4:**
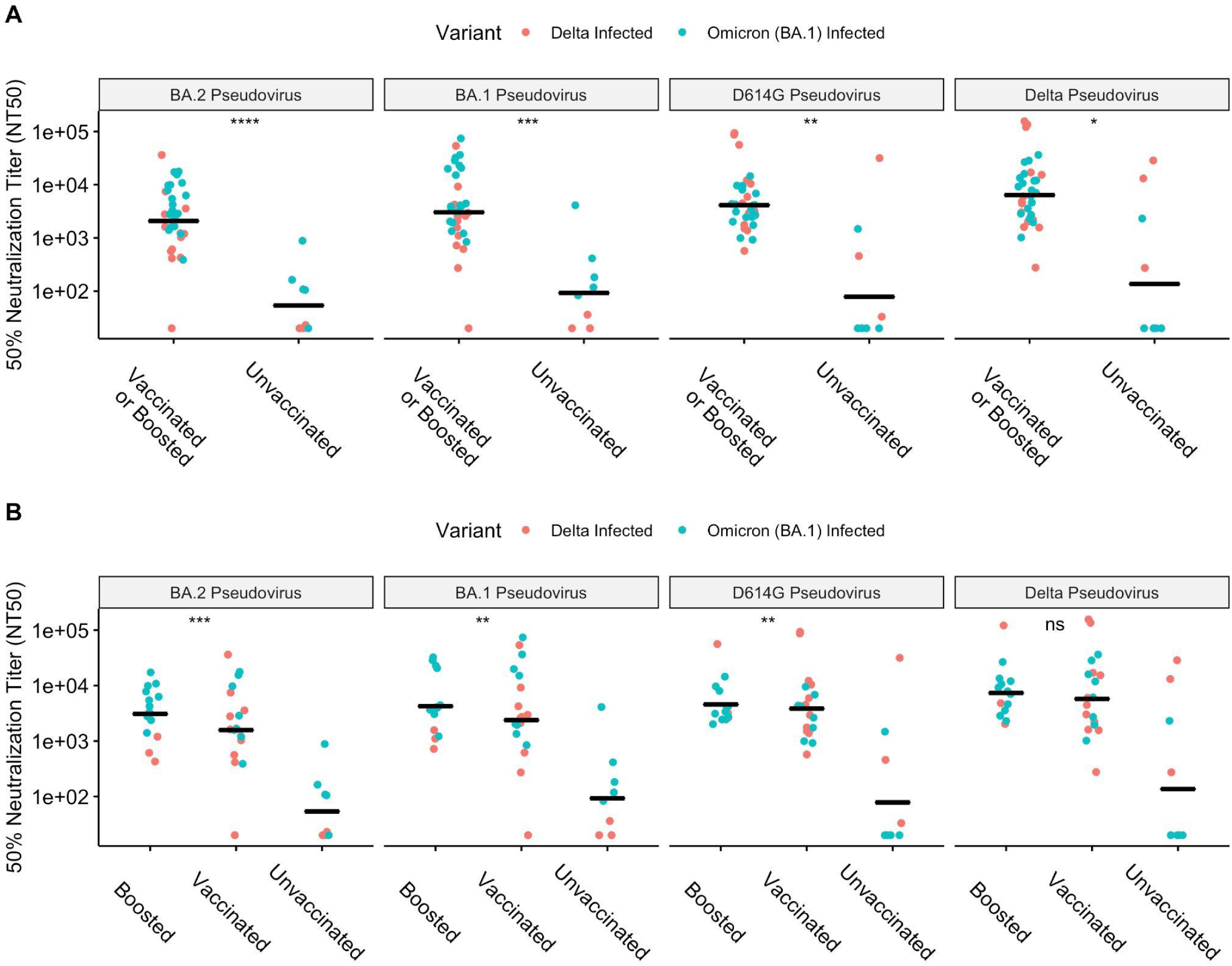
**A**. Neutralizing antibody responses, as measured by the 50% neutralizing antibody titer (NT50), at the time of convalescence, in unvaccinated, vaccinated or boosted individuals, against a panel of pseudoviruses. Significance from a Wilcoxon rank-sum test is shown. **B**. Neutralizing antibody responses, as measured by the 50% neutralizing antibody titer (NT50), at the time of convalescence, in unvaccinated, vaccinated, or boosted individuals, against a panel of pseudoviruses. Significance from a Kruskal-Wallis test is shown. ns: p > 0.05; * - p < 0.05; ** - p < 0.01; *** - p < 0.001; **** - p < 0.0001.

